# Levodopa Metabolism, Folate Deficiency, and Cardiovascular Risk in Lewy Body Dementia

**DOI:** 10.1101/2025.10.24.25338514

**Authors:** Yuji Higaki

## Abstract

**Background:** L-DOPA remains the cornerstone of Parkinson’s disease (PD) therapy, yet its long-term metabolic consequences are poorly understood. Folate deficiency appears disproportionately frequent in patients with dementia with Lewy bodies (DLB), a synucleinopathy closely related to PD. We hypothesized that folate deficiency is more prevalent in DLB than in Alzheimer’s disease (AD) or cognitively normal controls (NC), possibly due to excessive folate consumption during L-DOPA metabolism, which may elevate homocysteine levels and increase vascular risk.

**Methods:** We conducted a retrospective cross-sectional study comparing serum folate levels among DLB (n = 15), AD (n = 73), and NC (n = 7). Deficiency rates for folate, vitamin B12 (B12), and vitamin B1 (B1) were compared using Fisher’s exact test. Within DLB, associations) between folate/B12 deficiency and L-DOPA usage were assessed. Scatter plots visualized relationships between daily L-DOPA dosage and serum folate or B12 levels. Macrocytosis (MCV >100 fL) was evaluated as a potential indicator of vitamin depletion.

**Results:** Folate deficiency was significantly more prevalent in DLB than in AD or NC (p = 0.0021). Within DLB, folate deficiency was more frequent among L-DOPA users (p = 0.0406), while B12 deficiency showed no significant association. B12 levels varied widely, with some patients showing elevated concentrations. In contrast, folate levels tended to fall below the reference limit in all but one L-DOPA user. MCV was not a reliable indicator of deficiency.

**Conclusion:** L-DOPA–treated DLB patients may require individualized monitoring and tailored supplementation to mitigate vascular risk.

## Introduction

PD has emerged as a global health challenge, with its prevalence rising sharply in aging populations worldwide. [1, 2] Beyond its motor symptoms, PD is increasingly recognized for its association with cardiovascular events, [3–5] a concern consistently reported across epidemiological studies. [6] However, the underlying mechanisms contributing to this elevated cardiovascular risk remain poorly understood.

In our daily clinical practice, we have repeatedly observed that patients with DLB—a subtype of synucleinopathy [7, 8] closely related to PD—frequently present with folate deficiency. DLB, PD, and MSA are collectively referred to as synucleinopathies, characterized by pathological accumulation of alpha-synuclein in neurons or glial cells.^8^

In our clinical experience, folate deficiency in DLB is not an occasional finding but a recurring pattern. This observation raised a critical clinical question: Is folate deficiency statistically more prevalent in DLB patients compared to those with AD or NCIf so, what pathophysiological mechanisms might explain this discrepancy?

To address this question, we conducted a comparative study across three groups—DLB, AD, and NC—focusing on serum folate levels and their potential association with macrocytosis and cardiovascular vulnerability. Our aim was to clarify whether folate deficiency is disproportionately represented in DLB and to explore its implications for vascular risk in neurodegenerative disease.

## Materials and methods

The Institutional Review Board of the Yowa Hospital approved this study. The requirement for informed consent was waived because of the anonymous nature of the data.

### Study design and Setting

This observational study employed a retrospective cross-sectional design to analyze data from 133 consecutive patients who visited the memory clinic at Yowa Hospital between November 2020 and August 2025.

### Participants

Of the 133 patients who presented with memory complaints, 32 were diagnosed with conditions other than AD, DLB, or NC, and were excluded from the analysis. Among the remaining 101 patients, 6 AD cases lacked serum folate measurements and were excluded. These exclusions were deemed unlikely to affect the overall results. Consequently, 95 patients were included in the final analysis: 73 with AD, 15 with DLB, and 7 with NC (Fig. 1).

**Figure 1.**
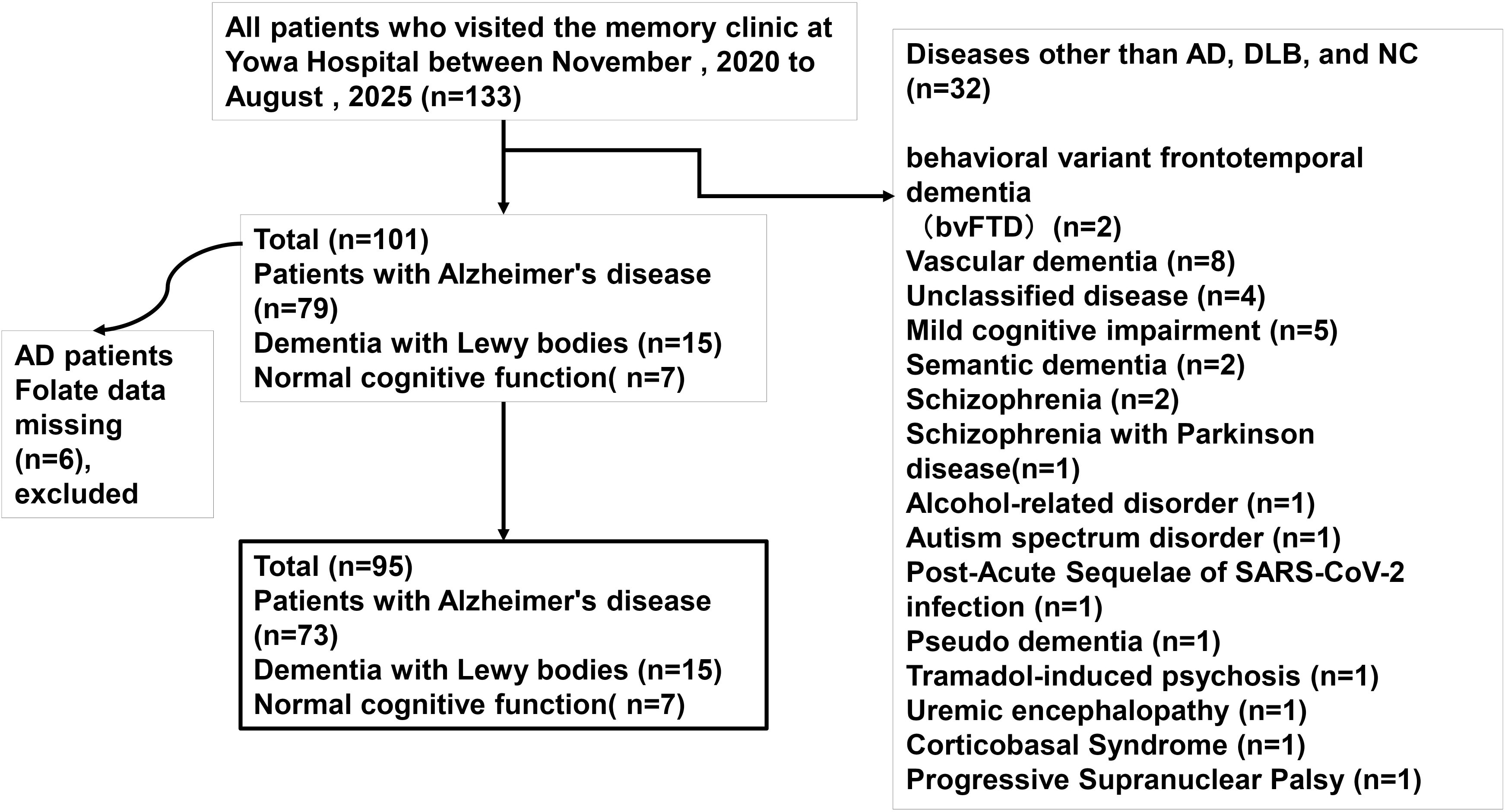
Flowchart of patient selection at Yowa Hospital memory clinic between November 2020 and August 2025. Of 133 initial patients, 95 were included in the final analysis after excluding those with missing folate data and non-target diagnoses.

### Variables and endpoints

All patients were personally evaluated by the author through clinical interviews, physical and neuropsychological examinations. Dementia was diagnosed according to According to the Diagnostic and Statistical Manual of Mental Disorders, Fifth Edition (DSM-5) criteria, and DLB was classified based on the consensus criteria proposed by McKeith et al. [9] The diagnostic process incorporated clinical symptoms, cognitive assessments, imaging findings, and laboratory results.

### Variables

Baseline variables included age, sex, years of education, Mini-Mental State Examination (MMSE) [10] score including the pentagon copying test (PCT). [11] The PCT, the final item of the MMSE, assesses visuospatial construction and has been independently studied as a sensitive marker for dementia subtypes.

And laboratory parameters: serum albumin, hemoglobin, red blood cell count, hematocrit, mean corpuscular volume (MCV), C-reactive protein (CRP), chronic kidney disease (CKD) status, serum iron, ferritin, and serum concentrations of B1, B12, and folate.

### Data sources/measurement

Age and years of education were obtained from medical records and patient interviews. Cognitive function was assessed using the MMSE, scored out of 30 points. A cut-off score of 24 is commonly used in previous studies to indicate cognitive impairment.

Laboratory parameters were measured using standardized automated assays at the hospital’s central laboratory. Continuous variables included serum albumin (g/dL), hemoglobin (Hb, g/dL), hematocrit (Ht, %), mean corpuscular volume (MCV, fL), hemoglobin A1c (HbA1c, NGSP, %), triglycerides (TG, mg/dL), low-density lipoprotein cholesterol (LDL-C, mg/dL), high-density lipoprotein cholesterol (HDL-C, mg/dL), and red blood cell count (RBC, ×10C/μL).

Categorical variables included sex and dichotomized laboratory values. MCV was categorized as ≤100.0 fL or >100.0 fL. CRP was considered negative if ≤0.3 mg/dL and positive if >0.3 mg/dL. CKD status was defined based on serum creatinine levels: ≥1.08 mg/dL for males and ≥0.80 mg/dL for females were considered positive; values below these thresholds were considered negative.

Vitamin deficiency rates were defined as follows: B1 deficiency was defined as serum B1 <2.6 μg/dL; B12 deficiency as serum B12 <180 pg/mL; and folate deficiency as serum folate <4.0 ng/mL. Deficiency rates were calculated as the proportion of deficient individuals within each diagnostic group: AD, DLB, and NC.

### Bias

For the MCV regression model—a secondary analysis not central to the study’s primary hypothesis—covariates were selected based on clinical plausibility and prior literature. To avoid overfitting, the number of predictors was limited in accordance with sample size considerations. While the conventional rule recommends one predictor per 15 cases, recent literature supports a more permissive threshold of one per 10 cases in exploratory clinical models. Variance inflation factors (VIFs) for all predictors were below 2.0, indicating minimal multicollinearity and stable model estimates.

### Study size

Sample size estimation using EZR (Ver.1.68), a free statistical software program with extended functions of R and R commander [12], indicated that 28 participants per group would be required to detect a difference in folate deficiency rates between the AD and DLB groups (5.5% vs. 40%) with 80% power and a two-sided alpha of 0.05. Although the DLB group included only 15 participants, a statistically significant difference was still observed (p = 0.0021). Post hoc power analysis using G*Power 3.1.9.7 [13] confirmed that the achieved statistical power was 0.893, indicating sufficient sensitivity to detect a meaningful difference.

### Endpoints and Statistical Analysis

The primary endpoints were the presence or absence of B1, B12, and folate deficiencies across the three diagnostic groups: AD, DLB, and NC. Group comparisons were performed using Fisher’s exact test.

Following this, a subgroup analysis was planned within the DLB cohort (n = 15), stratifying patients by L-DOPA usage. The presence of folate deficiency was compared between users and non-users using Fisher’s exact test.

To explore the relationship between L-DOPA dosage and vitamin status, scatter plots were generated within the DLB group to visualize associations between daily L-DOPA dose and serum concentrations of B12 and folate.

Among all 95 participants, the association between MCV and the presence of B12 or folate deficiency was evaluated using the independent samples t-test, as MCV values were confirmed to follow a normal distribution based on normality testing.

To identify predictors of MCV, multiple linear regression analysis was performed using MCV as the dependent variable and the following independent variables: serum albumin, B12 deficiency status, folate deficiency status, CRP positivity, CKD status, serum iron, and ferritin, Age.

Missing values for serum iron and ferritin (20 and 16 cases, respectively) were addressed using complete case analysis (CCA), resulting in a regression model based on 72 cases. Multiple imputation (MI) was considered but ultimately not adopted, as the inclusion of iron-related variables was deemed to dilute the study’s primary focus on vitamin-related predictors.

The phased expansion of laboratory testing at our memory clinic accounted for the missing values in serum iron and ferritin. These parameters were not included in routine assessments during the first year of the clinic’s operation (from November 2020), and were gradually incorporated based on literature review and clinical relevance.

### Ethical Considerations

All clinical procedures, including computed tomography (CT) imaging and blood sampling, were conducted as part of routine outpatient care. The use of these data for research purposes adhered to the ethical guidelines of Yowa Hospital. The study was approved by the hospital’s Ethics Committee (Approval No. 2020-02) and conducted in accordance with the Declaration of Helsinki (2013 revision).

## Results

### Participant Characteristics

Baseline characteristics are presented in Table 1.

**Table 1.** Baseline characteristics of study participants by diagnostic group. The cohort included 73 patients with Alzheimer’s disease (AD), 15 with dementia with Lewy bodies (DLB), and 7 cognitively normal controls. Variables are presented as mean (SD), median (IQR), or percentage, as appropriate. Subgroup comparisons by sex are also shown for hematological, metabolic, and cognitive parameters. Abbreviations: RBC = red blood cell count; Ht = hematocrit; MCV = mean corpuscular volume; CRP = C-reactive protein; CKD = chronic kidney disease; VB12 = vitamin B12; HbA1c = hemoglobin A1c; MMSE = Mini-Mental State Examination; LDL-C = low-density lipoprotein cholesterol; TG = triglycerides; HDL-C = high-density lipoprotein cholesterol.

**Baseline characteristics of study participants by diagnostic group**
| **variables** | **AD (n=73)** |  | **DLB (n=15)** |  | **Controls (n=7)** |  |
| --- | --- | --- | --- | --- | --- | --- |
| Age, year (SD) | 83.7 (6.7) |  | 83.4 (7.8) |  | 87.0 (6.1) |  |
| Sex, female (%) | 51 (69.9) |  | 13 (86.7) |  | 5 (71.4) |  |
| Years of education (IQR) | 12.0 (6.0-16.0) |  | 9.0 (6.0-12.0) |  | 12.0 (8.0-16.0) |  |
| Albumin, g/dl (IQR) | 4.1 (2.9-4.9) |  | 3.9 (2.8-5.0) |  | 3.9 (3.8-4.2) |  |
|  | AD Male (n=22) | AD Female (n=51) | DLB Male (n=2) | DLB Female (n=13) | Controls Male (n=2) | Controls Female (n=5) |
| Hemoglobin, g/dl (SD) | 13.3 (1.6) | 12.9 (1.1) | 12.8 (4.0) | 12.1 (1.1) | 13.3 (2.1) | 12.5 (0.5) |
| RBC ( × 10<sup>6</sup>/μL) (SD) | 4.23 (0.4) | 4.23 (0.4) | 4.10 (1.4) | 3.92 (0.4) | 4.34 (0.39) | 4.18 (0.25) |
| Ht (%) (SD) | 40.2 (4.6) | 39.1 (3.3) | 37.9 (13.8) | 35.9 (4.6) | 39.5 (4.6) | 37.7 (1.9) |
| MCV ( f l ) | 93.4 (4.4) |  | 93.1 (3.6) |  | 90.6 (4.7) |  |
| MCV.≤99.9 (%) | 67 (91.8) |  | 15 (100.0) |  | 7 (100.0) |  |
| MCV.≥100 (%) | 6 (8.2) |  | 0 (0.0) |  | 0 (0.0) |  |
| CRP positive (%) | 12 (16.4) |  | 2 (13.3) |  | 0 (0.0) |  |
| CRP negative (%) | 61 (83.6) |  | 13 (86.7) |  | 7 (100.0) |  |
| CKD positive (%) | 26 (35.6) |  | 6 (40.0) |  | 2 (28.6) |  |
| CKD negative (%) | 47 (64.4) |  | 9 (60.0) |  | 5 (71.4) |  |
| VB1.def (%) | 18 (24.7) |  | 4 (26.7) |  | 2 (28.6) |  |
| VB12.def (%) | 15 (20.5) |  | 1 (6.7) |  | 1 (14.3) |  |
| folate def (%) | 4 (5.5) |  | 6 (40.0) |  | 0 (0.0) |  |
| HbA1c (NGSP)(%) | 5.6 (4.7-10.5) |  | 5.4 (4.8-8.1) |  | 5.7 (5.0-7.0) |  |
| MMSE score, points | 19.8 (5.5) |  | 20.3 (4.7) |  | 28.8 (1.8) |  |
| Pentagon copying failure (%) | 23 (32.9) |  | 8 (61.5) |  | 0 (0.0) |  |
| LDL-C (mg/dl) | 116 (59-201.0) |  | 116 (68-239.0) |  | 111 (86.0-167.0) |  |
|  | AD Male (n=22) | AD Female (n=51) | DLB Male (n=2) | DLB Female (n=13) | Controls Male (n=2) | Controls Female (n=5) |
| TG (mg/dl) | 90.5 (67.3-132.8) | 99.0 (73.5-153.0) | 77.5 (55.9) | 104.3 (32.2) | 154 (18.4) | 143 (57.5) |
| HDL-C (mg/dl) | 59.5 (47.3-66.0) | 74.0 (63.5-88.5) | 71.0 (5.7) | 70.2 (20.7) | 57.5 (21.9) | 70.6 (6.6) |

No significant differences were observed in age, sex, albumin, or Hb levels across groups. Years of education were significantly lower in the DLB group (p = 0.032). MMSE scores were significantly reduced in AD and DLB compared to NC (p = 0.001), and pentagon copying failure was more frequent in DLB than AD (p = 0.025).

### Vitamin Deficiency

Folate deficiency was observed in 40.0% of DLB patients, significantly higher than in AD (5.5%) and NC (0.0%) (p = 0.0021). No significant differences were found in B1 (p = 1.0) or B12 deficiency rates (p = 0.51).

These associations are summarized in Table 2.

**Table 2.** Frequency of vitamin deficiency across diagnostic groups (Alzheimer’s disease [AD], dementia with Lewy bodies [DLB], and cognitively normal controls [NC]). Deficiency status for vitamin B1, vitamin B12, and folate was compared using Fisher’s exact test. Folate deficiency was significantly more frequent in the DLB group (p = 0.0021), while no significant differences were observed for vitamin B1 (p = 1.0) or vitamin B12 (p = 0.51). Deficiency was defined according to laboratory reference ranges. *p < 0.05.

**Fisher's exact test for vitamin deficiency across diagnostic groups (AD, DLB, NC).**
|  | AD | DLB | NC | p value |
| --- | --- | --- | --- | --- |
| VB1. def. | 18 | 4 | 2 | p=1.0 |
| VB1 Normal | 55 | 11 | 5 |  |
| VB12. def | 15 | 1 | 1 | p=0.51 |
| VB12,. Normal | 58 | 14 | 6 |  |
| Folate. def | 4 | 6 | 0 | p=0.0021* |
| Folate. Normal | 69 | 9 | 7 |  |

Note: Deficiency was defined based on laboratory reference ranges. P-values were calculated using Fisher's exact test.
\*p<0.05
AD: Alzheimer's disease; DLB: dementia with Lewy bodies; NC: normal cognition.

### L-DOPA and Folate Status

Among patients with DLB, folate deficiency was significantly more frequent in those receiving L-DOPA (5/7) compared to those not receiving L-DOPA (1/8) (p = 0.0406). No significant association was found between L-DOPA use and B12 deficiency (p = 1.0). These findings are summarized in Table 3.

**Table 3.** Association between L-DOPA use and folate or vitamin B12 deficiency in patients with dementia with Lewy bodies (DLB, n = 15). Deficiency status was compared between L-DOPA users and non-users using Fisher’s exact test. Folate deficiency was significantly more frequent among L-DOPA users (p = 0.0406), while no significant association was found for vitamin B12 deficiency (p = 1.0). Deficiency was defined according to laboratory reference ranges. *p < 0.05.

**Association between L-DOPA use and folate or vitamin B12 deficiency in patients with DLB (n = 15).**
|  | folate-def. | folate-Normal |  |
| --- | --- | --- | --- |
| L-DOPA(-) | 1 | 7 | P = 0.0406* |
| L-DOPA(+) | 5 | 2 |  |

|  | VB12-def. | VB12-Normal |  |
| --- | --- | --- | --- |
| L-DOPA(-) | 1 | 7 | P = 1 |
| L-DOPA(+) | 0 | 7 |  |

Note: Deficiency was defined based on laboratory reference ranges. P-values were calculated using Fisher's exact test.
\*p<0.05
DLB: dementia with Lewy bodies

Fig. 2 illustrates these relationships, showing scatter plots of daily L-DOPA dosage and serum folate (A) and B12 (B) concentrations.

**Figure 2.**
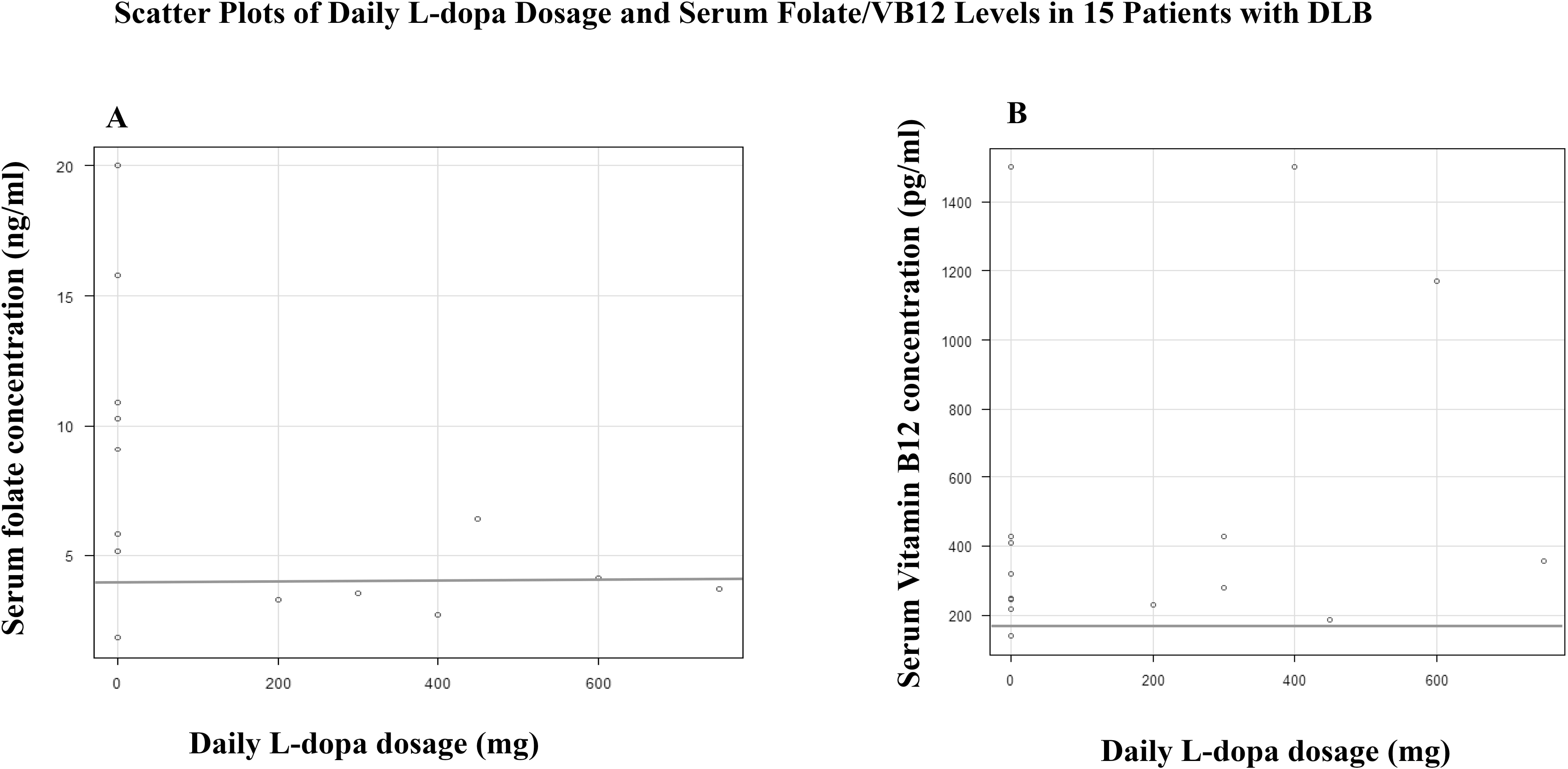
Scatter plots of daily L-DOPA dosage and serum vitamin concentrations in 15 patients with dementia with Lewy bodies (DLB). (A) Serum folate levels show a dose-dependent decline with increasing L-DOPA dosage, suggesting metabolic depletion. (B) Serum vitamin B12 levels remain scattered and unaffected by dosage, indicating no dose-dependent association. Overlapping data points are annotated in the figure for visual clarity.

Folate concentrations were predominantly below reference thresholds in L-DOPA users, while B12 levels remained within normal limits regardless of dosage. Although each scatter plot represents data from 15 patients, only 14 dots are visually distinguishable in each figure due to overlapping values. In the B12 plot, two patients had similar serum levels (245 and 247 pg/ml), resulting in near-complete overlap. In the folate plot, two patients shared an identical value of 3.5 ng/ml, leading to complete dot overlap.

### MCV

MCV did not differ significantly between participants with and without B12 deficiency (92.2CfL (5.7) vs. 93.4CfL (4.0), p = 0.31) or between those with and without folate deficiency (92.1CfL (4.1) vs. 93.3CFl (4.4), p = 0.41), as determined by independent samples t-tests.

Only 6 out of 95 participants (6.3%) had an MCV greater than 100 fL, indicating that macrocytosis was rare across the cohort. [15]

These comparisons are illustrated in Fig. 3.

**Figure 3.**
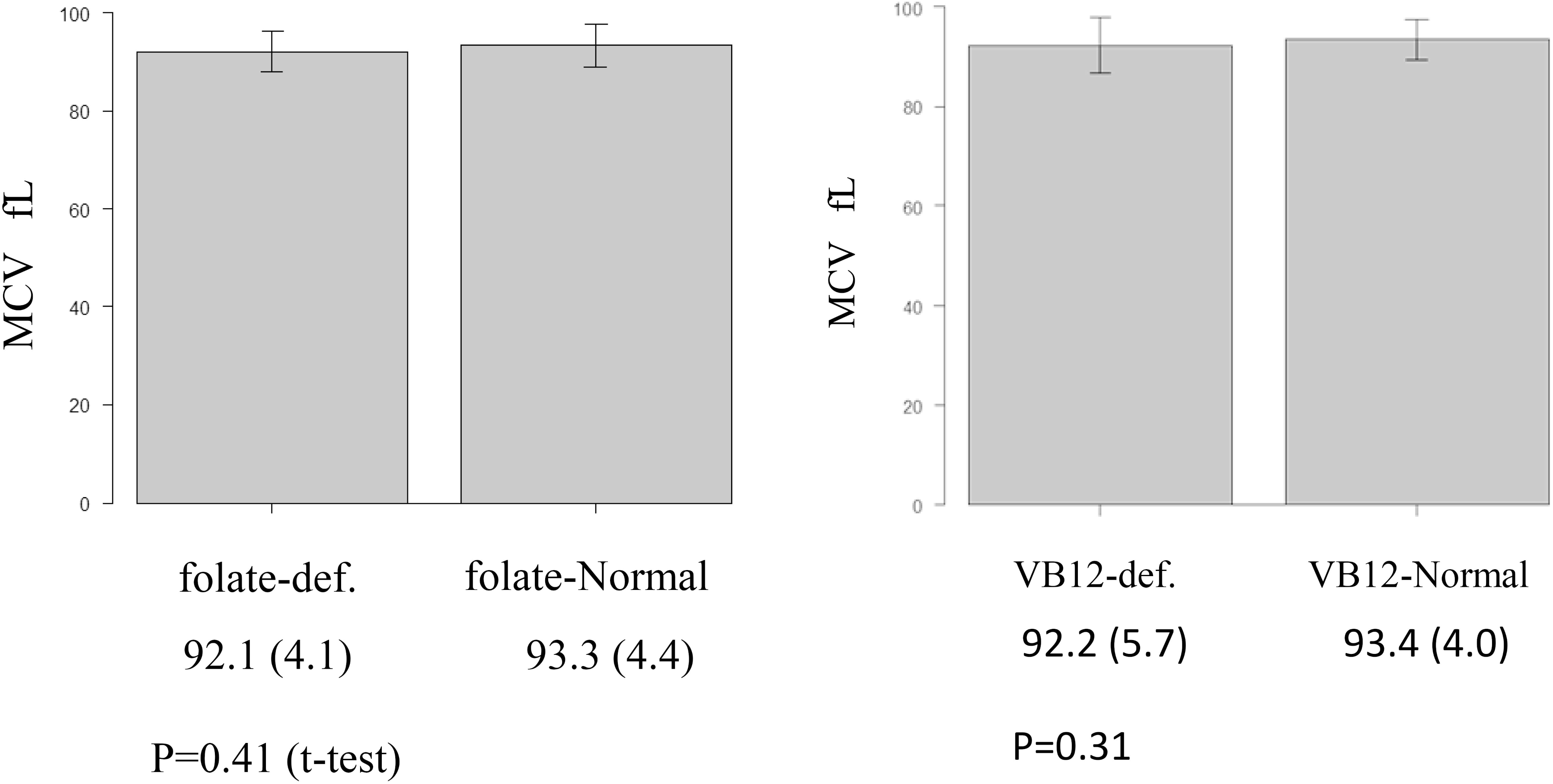
Comparison of mean corpuscular volume (MCV) between vitamin-deficient and non-deficient groups among 95 participants (AD: 73, DLB: 15, NC: 7). (A) MCV values in vitamin B12-deficient vs. B12-normal groups: 92.2 fL (5.7) vs. 93.4 fL (4.0), p = 0.31. (B) MCV values in folate-deficient vs. folate-normal groups: 92.1 fL (4.1) vs. 93.3 fL (4.4), p = 0.41. Group comparisons were performed using independent samples t-tests, following confirmation of normality. No statistically significant differences were observed, indicating that MCV was not a reliable indicator of vitamin deficiency in this cohort.

### Predictors of MCV

Multiple linear regression analysis was performed using complete case data (n = 72) to identify predictors of MCV. Among the eight variables tested, serum iron (unstandardized coefficient = 0.050, 95% CI: 0.009–0.091, p = 0.018) and CRP positivity (unstandardized coefficient = 2.91, 95% CI: 0.017–5.80, p = 0.049) were significantly associated with MCV.

In contrast, folate deficiency (p = 0.24) and B12 deficiency (p = 0.75) were not significantly associated with MCV variation. These findings suggest that macrocytic changes in this cohort were more closely linked to iron metabolism and inflammatory status than to vitamin deficiency.

A summary of the regression coefficients and confidence intervals is presented in Fig. 4.

**Figure 4.**
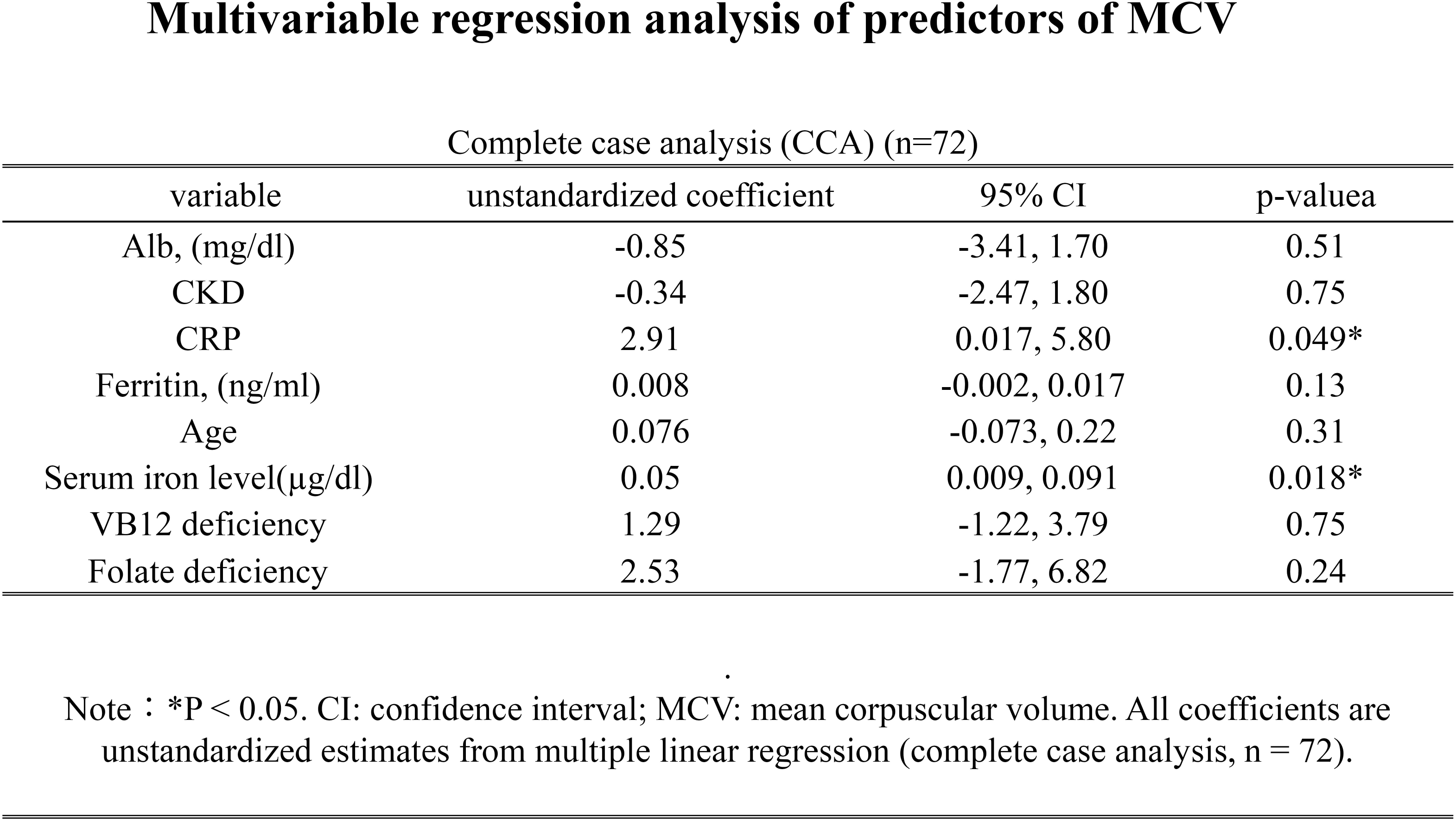
Results of multiple linear regression analysis examining predictors of mean corpuscular volume (MCV) in a complete case cohort (n = 72). Eight independent variables were tested: serum albumin, chronic kidney disease (CKD) status, C-reactive protein (CRP) positivity, serum ferritin, serum iron, age, and deficiency status of vitamin B12 and folate. Serum iron (unstandardized coefficient = 0.050, 95% CI: 0.009–0.091, p = 0.018) and CRP positivity (unstandardized coefficient = 2.91, 95% CI: 0.017–5.80, p = 0.049) were significantly associated with MCV. No significant associations were observed for albumin, CKD, ferritin, age, or vitamin deficiency status.

## Discussion

We conducted an observational retrospective cross-sectional study involving 95 patients (AD: n=73, DLB: n=15, NC: n=7). The key findings were as follows: (1) Folate deficiency was significantly more prevalent in the DLB group (40.0%) compared to AD (5.5%) and NC (0.0%). (2) Within the DLB cohort, folate deficiency was more frequent among patients receiving L-DOPA, whereas no association was found with B12 deficiency. (3) Scatterplot analysis revealed a dose-dependent decline in serum folate levels with increasing daily L-DOPA dosage, while B12 levels remained scattered and unaffected. (4) MCV failed to serve as a reliable indicator of either folate or B12 deficiency.[14, 15]

Previous studies have consistently reported elevated rates of ischemic heart disease and stroke among patients with parkinsonism (including PD and DLB). A 20-year prospective cohort study further confirmed these findings, demonstrating significantly higher vascular risk compared to age-matched cognitively normal controls. [17]

However, recent reviews have largely emphasized autonomic dysfunction as a primary mechanism, leaving the vascular pathogenesis unresolved. [3, 17] Our findings suggest that folate—not B12—may be preferentially depleted in L-DOPA-treated patients, potentially contributing to vascular vulnerability. This hypothesis is supported by Clarke’s 2000 report, [6] which demonstrated a rise in standardized mortality rates among PD patients in Wales beginning around 1980—approximately a decade after the widespread adoption of L-DOPA therapy. Given that homocysteine, a pro-atherogenic metabolite of L-DOPA metabolism, requires folate [17] and B12 for detoxification, chronic folate depletion may plausibly accelerate vascular pathology over time. [18]

Unexpectedly, multiple regression analysis revealed no significant association between MCV and either folate or B12 deficiency. [14, 15] Instead, CRP positivity and serum iron levels emerged as independent predictors of MCV variation. These findings underscore the limited utility of MCV in detecting vitamin deficiency in PD/DLB patients and highlight the need for direct measurement of serum folate and B12. While optimal testing intervals remain to be determined, we encourage clinicians worldwide to validate and replicate these observations. Given the established role of homocysteine in vascular disease, we believe that daily supplementation with folate and B12 may offer a pragmatic and ethically sound strategy to mitigate cardiovascular risk in L-DOPA-treated patients. [19]

### Limitations

This study has several limitations.

First, homocysteine—a key biomarker in folate metabolism and vascular risk—was not measured. Although clinically relevant, homocysteine testing is not routinely performed at our institution due to reimbursement constraints. In the current healthcare climate in Japan, where hospital management faces increasing financial scrutiny, certain laboratory tests—particularly outsourced ones—are selectively ordered to avoid potential insurance audits. As homocysteine testing may be subject to such scrutiny, it was excluded from our protocol.

Second, serum iron and ferritin values were missing in a subset of patients due to the phased expansion of laboratory testing at our memory clinic. These parameters were not included in routine assessments during the clinic’s first year of operation. We employed CCA for regression modeling, focusing on a single model to examine predictors of MCV.

Third, the sample size of the DLB group was relatively small. However, the observed difference in folate deficiency rates between DLB and AD groups reached statistical significance, and post hoc power analysis confirmed adequate sensitivity to detect meaningful effects.

Lastly, as a cross-sectional study, causal relationships cannot be inferred. Longitudinal studies incorporating homocysteine measurements and larger DLB cohorts are warranted to further elucidate the mechanistic link between L-DOPA metabolism, folate depletion, and cardiovascular risk.

In conclusion, this study demonstrates that folate deficiency is significantly more prevalent in patients with DLB than in those with AD or cognitively normal controls. Within the DLB cohort, folate deficiency was strongly associated with L-DOPA usage, and serum folate levels declined in a dose-dependent manner with increasing daily L-DOPA dosage. In contrast, B12 levels remained unaffected, suggesting a selective metabolic vulnerability of folate in this population.

## Data Availability

All raw clinical data used in this study are available upon reasonable request, with appropriate anonymization and subject to approval by the director of the hospital where the study was conducted.

## Acknowledgment

The author gratefully acknowledges Mr. Hiroki Yoshida, MSc, founder of DataSeed Inc., for his expert statistical advice. He is currently pursuing doctoral studies and has extensive experience in pharmaceutical and clinical research settings.

The patient selection process is illustrated in Fig. 1.

## References

1. Tolosa E, Garrido A, Scholz SW, Poewe W. Challenges in the diagnosis of Parkinson’s disease. Lancet Neurol. 2021 May;20(5):385–397. doi: 10.1016/S1474-4422(21)00030-2. PMID: 33894193; PMCID: PMC8185633.

2. Jankovic J, Tan EK. Parkinson’s disease: etiopathogenesis and treatment. J Neurol Neurosurg Psychiatry. 2020 Aug;91(8):795–808. doi: 10.1136/jnnp-2019-322338. Epub 2020 Jun 23. PMID: 32576618.

3. Grosu L, Grosu AI, Crisan D, Zlibut A, Perju-Dumbrava L. Parkinson’s disease and cardiovascular involvement: Edifying insights (Review). Biomed Rep. 2023 Feb 14;18(3):25. doi: 10.3892/br.2023.1607. PMID: 36846617; PMCID: PMC9944619.

4. Rogers JD, Sanchez-Saffon A, Frol AB, Diaz-Arrastia R. Elevated plasma homocysteine levels in patients treated with levodopa: association with vascular disease. Arch Neurol. 2003 Jan;60(1):59–64. doi: 10.1001/archneur.60.1.59. PMID: 12533089.

5. Günaydın ZY, Özer FF, Karagöz A, Bektaş O, Karataş MB, Vural A, Bayramoğlu A, Çelik A, Yaman M. Evaluation of cardiovascular risk in patients with Parkinson disease under levodopa treatment. J Geriatr Cardiol. 2016 Jan;13(1):75–80. doi: 10.11909/j.issn.1671-5411.2016.01.003. PMID: 26918017; PMCID: PMC4753016.

6. CLarke CE. Mortality from Parkinson’s disease. J Neurol Neurosurg Psychiatry. 2000 Feb;68(2):254–5. doi: 10.1136/jnnp.68.2.254. PMID: 10702045; PMCID: PMC1736768.

7. Jellinger KA, Korczyn AD. Are dementia with Lewy bodies and Parkinson’s disease dementia the same disease? BMC Med. 2018 Mar 6;16(1):34. doi: 10.1186/s12916-018-1016-8. PMID: 29510692; PMCID: PMC5840831.

8. Uchihara T, Giasson BI. Propagation of alpha-synuclein pathology: hypotheses, discoveries, and yet unresolved questions from experimental and human brain studies. Acta Neuropathol. 2016 Jan;131(1):49–73. doi: 10.1007/s00401-015-1485-1. Epub 2015 Oct 7. PMID: 26446103; PMCID: PMC4698305.

9. McKeith IG, Boeve BF, Dickson DW, Halliday G, Taylor JP, Weintraub D, Aarsland D, Galvin J, Attems J, Ballard CG, Bayston A, Beach TG, Blanc F, Bohnen N, Bonanni L, Bras J, Brundin P, Burn D, Chen-Plotkin A, Duda JE, El-Agnaf O, Feldman H, Ferman TJ, Ffytche D, Fujishiro H, Galasko D, Goldman JG, Gomperts SN, Graff-Radford NR, Honig LS, Iranzo A, Kantarci K, Kaufer D, Kukull W, Lee VMY, Leverenz JB, Lewis S, Lippa C, Lunde A, Masellis M, Masliah E, McLean P, Mollenhauer B, Montine TJ, Moreno E, Mori E, Murray M, O’Brien JT, Orimo S, Postuma RB, Ramaswamy S, Ross OA, Salmon DP, Singleton A, Taylor A, Thomas A, Tiraboschi P, Toledo JB, Trojanowski JQ, Tsuang D, Walker Z, Yamada M, Kosaka K. Diagnosis and management of dementia with Lewy bodies: Fourth consensus report of the DLB Consortium. Neurology. 2017 Jul 4;89(1):88–100. doi: 10.1212/WNL.0000000000004058. Epub 2017 Jun 7. PMID: 28592453; PMCID: PMC5496518.

10. Folstein MF, Folstein SE, McHugh PR. “Mini-mental state”. A practical method for grading the cognitive state of patients for the clinician. J Psychiatr Res. 1975 Nov;12(3):189–98. doi: 10.1016/0022-3956(75)90026-6. PMID: 1202204.

11. Kim N, Truty T, Han SD, Heo M, Buchman AS, Bennett DA, Tasaki S. Digital quantification of the MMSE interlocking pentagon areas: a three-stage algorithm. Sci Rep. 2024;14:59194. doi:10.1038/s41598-024-59194-1

12. Kanda Y. Investigation of the freely available easy-to-use software ‘EZR’ for medical statistics. Bone Marrow Transplant. 2013 Mar;48(3):452–8. doi: 10.1038/bmt.2012.244. Epub 2012 Dec 3. PMID: 23208313; PMCID: PMC3590441.

13. Faul F, Erdfelder E, Buchner A, Lang AG. Statistical power analyses using G*Power 3.1: tests for correlation and regression analyses. Behav Res Methods. 2009 Nov;41(4):1149–60. doi: 10.3758/BRM.41.4.1149. PMID: 19897823.

14. Nagao T, Hirokawa M. Diagnosis and treatment of macrocytic anemias in adults. J Gen Fam Med. 2017 Apr 13;18(5):200–204. doi: 10.1002/jgf2.31. PMID: 29264027; PMCID: PMC5689413.

15. Bando T, Tokuda M, Katsuda I, Emi N, Tomita A. Involvement of folate and vitamin B12 deficiency in patients with normocytic anemia. Fujita Med J. 2023 May;9(2):134–141. doi: 10.20407/fmj.2022-016. Epub 2022 Oct 28. PMID: 37234385; PMCID: PMC10206897.

16. Ben-Shlomo Y, Marmot MG. Survival and cause of death in a cohort of patients with parkinsonism: possible clues to aetiology? J Neurol Neurosurg Psychiatry. 1995 Mar;58(3):293–9. doi: 10.1136/jnnp.58.3.293. PMID: 7897409; PMCID: PMC1073364.

17. Chua SKK, Saffari SE, Lee SJY, Tan EK. Association Between Parkinson’s Disease and Coronary Artery Disease: A Systematic Review and Meta-Analysis. J Parkinsons Dis. 2022; 12(6):1737–1748. doi: 10.3233/JPD-223291. PMID: 35694936; PMCID: PMC9789484.

18. Voutilainen S, Lakka TA, Porkkala-Sarataho E, Rissanen T, Kaplan GA, Salonen JT. Low serum folate concentrations are associated with an excess incidence of acute coronary events: the Kuopio Ischaemic Heart Disease Risk Factor Study. Eur J Clin Nutr. 2000 May;54(5):424–8. doi: 10.1038/sj.ejcn.1600991. PMID: 10822291.

19. Anamnart C, Kitjarak R. Effects of vitamin B12, folate, and entacapone on homocysteine levels in levodopa-treated Parkinson’s disease patients: A randomized controlled study. J Clin Neurosci. 2021 Jun;88:226–231. doi: 10.1016/j.jocn.2021.03.047. Epub 2021 Apr 20. PMID: 33992189.

